# Age- and Gender-Specific Distribution and Susceptibility-Guided Multidrug Strategies in NTM Lung Disease: A retrospective study from a specialized hospital in China

**DOI:** 10.1101/2025.09.11.25335602

**Authors:** Hengjian Fan, Li Xu, Xiao Ru

**Author notes:** (X. Ru).

## Abstract

**Introduction:** Nontuberculous mycobacteria (NTM) are causative pathogens of NTM pulmonary disease (NTM-PD), which is managed with multidrug regimens guided by antimicrobial susceptibility testing (AST). This study evaluated the AST profiles of clinical NTM isolates from a specialized hospital and proposes targeted therapeutic strategies for NTM-PD. Methodology: Diagnosis of 606 NTM-PD cases complied with American Thoracic Society (ATS) criteria. Broth microdilution methodology quantified minimum inhibitory concentrations (MICs) for antimicrobial agents against NTM isolates obtained from sputum or bronchoalveolar lavage specimens. Susceptibility profiles among NTM species were systematically evaluated to establish optimized treatment regimens using pharmacodynamically validated breakpoint parameters.

**Results:** Epidemiological analysis revealed a statistically significant gender disparity in NTM-PD prevalence, with male predominance (male 54.79% *vs* female 45.21%) and increasing incidence correlating with advanced age. A total of 606 isolates were identified as 21 NTM species, predominantly *Mycobacterium avium* complex (MAC) (73.1%), *Mycobacterium abscessus* complex (MABC) (12.4%), and *Mycobacterium kansasii* (*M. kansasii*) (8.1%). MAC showed > 95% susceptibility to rifamycins, clarithromycin, amikacin, and linezolid, with resistance to imipenem/tetracyclines. MABC maintained > 80% susceptibility to linezolid, amikacin, clarithromycin, cefixime, and moxifloxacin but imipenem resistance. *M. kansasii* demonstrated 30.6% sulfamethoxazole susceptibility versus > 80% for clarithromycin, amikacin, fluoroquinolones, and linezolid. Optimal combinations: rifamycin-clarithromycin-linezolid (MAC) and linezolid-amikacin-ethambutol (MABC).

**Conclusions:** NTM species exhibit distinct antimicrobial profiles, with clarithromycin, linezolid, and amikacin demonstrating efficacy versus limited activity of imipenem/ sulfamethoxazole. Pulmonary NTM infections necessitate species-specific multidrug regimens guided by AST, while regional susceptibility epidemiology informs evidence-based empirical therapy and controlled trial frameworks.

## Introduction

NTM comprise over 200 recognized species and subspecies as of 2025 (http://www.bacterio.net/mycobacterium.html), with most being environmental organisms and only a subset causing human disease. Pulmonary infections account for most of NTM-related disorders, while other manifestations include cutaneous, osteoarticular, and neurological infections. Taxonomically classified under the family *Mycobacteriaceae* in the order *Actinomycetales*, the majority of NTM species demonstrate opportunistic pathogenicity. Clinically, NTM-PD represents the principal disease manifestation among human infections[1]. Historically, technological limitations in mycobacterial isolation and identification led to frequent misdiagnoses of NTM-PD as tuberculosis, complicating clinical management. Enhanced clinical awareness among respiratory specialists, coupled with advancements in molecular identification techniques and improved culture isolation protocols, has enabled more precise differentiation of NTM species from other acid-fast bacilli. Concurrent increases in host susceptibility factors have contributed to improved detection and diagnosis of NTM-associated diseases. Recent epidemiological studies document rising prevalence rates of NTM infections across multiple geographical regions, including Asia, North America, Europe, and Oceania[2]. This emerging pattern highlights the growing clinical relevance of NTM in modern medical practice.

Diagnosing NTM-PD necessitates a comprehensive assessment of clinical symptoms, radiological findings, and microbiological evidence, with culture-based identification serving as the diagnostic gold standard. Clinically[3], the management of NTM-PD presents multiple challenges. While the standard MAC triple therapy (clarithromycin or azithromycin + ethambutol + rifabutin) achieves sputum culture conversion in 50%–70% of cases[4], recurrence rates remain substantial. Treatment failure escalates to 25%–60% in cavitary or advanced disease[5]. For MABC infections, clinical response rates range from 30% to 50%, contrasting with > 90% efficacy observed in *M. kansasii* infections[4]. Current clinical guidelines issued by major thoracic societies, including the British Thoracic Society and ATS[3,6], along with specialized recommendations for rare NTM infections[7], uniformly advocate for AST following culture isolation. These consensus documents emphasize the necessity of tailoring therapeutic regimens according to AST results to optimize treatment outcomes, particularly given the inherent antibiotic resistance patterns observed in NTM species. However, key challenges persist. Limited evidence supports the use of certain drugs with promising in vitro activity, such as moxifloxacin and linezolid[8]. Standard regimens often require multi-drug combinations, leading to prolonged treatment durations and increased toxicity, highlighting the urgent need for optimized treatment strategies.

## Materials and methods

### Study subjects

This study analyzed 606 nontuberculous mycobacterial (NTM) isolates obtained from sputum or bronchoalveolar lavage fluid specimens at Shandong Provincial Public Health Clinical Center between January 2021 and October 2024. Duplicate isolates from individual patients were systematically excluded. All isolates originated from patients meeting the 2020 ATS diagnostic criteria for NTM pulmonary disease (NTM-PD). Comprehensive AST was conducted on all eligible isolates. Data were de-identified by removing all direct identifiers (e.g., patient names, medical record numbers) prior to processing, and the de-identified data were subsequently used for analysis.

### Experimental methods

Colloidal gold immunochromatographic assay was applied for antigen detection on BACTEC MGIT 960 culture-positive isolates, with subsequent species identification was implemented via DNA microarray analysis integrated with matrix-assisted laser desorption/ionization time-of-flight mass spectrometry. Antimicrobial susceptibility of selected nontuberculous mycobacterial (NTM) strains to 15 therapeutic agents was assessed via Broth microdilution methodology. Bacterial ultrasonic turbidity quantification was carried out using an ultrasonic dispersion counter, and the susceptibility interpretations are made in accordance with the standardized microplate assay protocols. CLSI documents M62-Ed1 and M24-Ed3 served as interpretive criteria[9,10], with species-specific clinical breakpoints for antimicrobial agents established based on MIC distribution patterns observed in Chinese domestic clinical isolates.

Species-specific clinical breakpoints for susceptibility interpretation were defined as follows: Clinical breakpoints for AST of MAC isolates: linezolid 8–32 µg/ml; clarithromycin 0.5–16 µg/ml; rifampicin 1–6µg/ml; amikacin 4–64 µg/ml; imipenem 4–16 µg/ml; ethambutol 1.25–5 µg/ml; ceftazidime 64–80 µg/ml; azithromycin 1–16 µg/ml; rifabutin 0.5–2 µg/ml; tobramycin 4–16 µg/ml; gatifloxacin 0.25–1 µg/ml; moxifloxacin 1–4 µg/ml; doxycycline 1–8 µg/ml; minocycline 1–8 µg/ml; sulfamethoxazole 32–80 µg/ml.

Clinical breakpoints for AST of MABC isolates: linezolid 8–32 µg/ml; clarithromycin 2–8 µg/ml; amikacin 16–64µg/ml; imipenem 1–4 µg/ml; ceftazidime 16–128 µg/ml; tobramycin 2–8 µg/ml; moxifloxacin 1–4 µg/ml; doxycycline 1–8 µg/ml; minocycline 1–8 µg/ml; sulfamethoxazole 32–80 µg/ml; azithromycin 1–16 µg/ml.

Clinical breakpoints for AST of *M. kansasii* isolates: linezolid 8–32 µg/ml; clarithromycin 0.5–16 µg/ml; rifampicin 1 µg/ml (susceptibility breakpoint); amikacin 32 µg/ml (susceptibility breakpoint); ethambutol 4 µg/ml (susceptibility breakpoint); rifabutin 0.5–2 µg/ml; gatifloxacin 2–8 µg/ml; moxifloxacin 2 µg/ml (susceptibility breakpoint); sulfamethoxazole 32–80 µg/ml.

Clinical breakpoints for AST of *Mycobacterium chelonei* (*M. chelonei*) and *M. fortuitum* isolates: the same as MABC.

Instruments and Reagents: BacT/ALERT-3D 480 mycobacterial detection system Manufacturer: bioMérieux, France. Roche LightCycler 480 real-time PCR system Manufacturer: Roche Diagnostics, Switzerland. LuxScan10K/B microarray scanner Manufacturer: CapitalBio Corporation, China. Bruker MALDI-TOF mass spectrometer Manufacturer: Bruker Daltonics, USA. BACspreader1100 Bacterial ultrasonic dispersion counter Manufacturer: Hangzhou Aosheng Instruments Co., Ltd., China. MPT64 antigen detection kit Manufacturer: Hangzhou Innovation Biotechnology Control Technology Co., Ltd., China. NTM drug susceptibility plate Manufacturer: Zhuhai Yinke Biotechnology Co., Ltd., China.

Conduct statistical analysis of demographic covariates (age and sex) in patients with NTM-PD. Statistical analyses were conducted to evaluate susceptibility patterns of NTM clinical isolates. Overall susceptibility of drug combinations was systematically assessed through in vitro susceptibility testing. Correlation analyses were conducted to determine resistance associations among structurally similar antimicrobial agents.

### Statistical analysis

Statistical analyses were conducted using SPSS 26 (IBM Corp.) and Microsoft Excel. In antimicrobial susceptibility testing for NTM, comparative analysis of susceptibility rates and inter-drug correlations within the same category requires selection of appropriate statistical methods based on sample size and expected frequency distribution parameters. The methodological framework is established as follows:1. For datasets with *n* ≥ 40 where all cells have expected frequencies ≥ 5, or when *n* ≥ 40 with ≤ 20% of cells having expected frequencies between 1 and 5, the Pearson chi-square test (Pearson *χ*² test) is appropriate; For 2×2 contingency tables with small sample sizes (*n* < 40), or when > 20% of cells exhibit expected frequencies of 1–5 with at least one cell < 1, Fisher’s exact test should be applied; 3. When data violate assumptions of Pearson *χ*² or Fisher’s exact test, the Monte Carlo simulation chi-square test (MC *χ*² test) is used, followed by calculation of Cramér’s V and Cohen’s kappa coefficients (Cohen’s *κ*). Cramér’s V interpretation thresholds were defined as: < 0.30 (weak association), 0.30–0.60 (moderate association), and > 0.60 (strong association). Cohen’s κ values were interpreted according to standardized criteria: values below zero indicated below-chance agreement (statistical non-significance), 0.00–0.20 denoted slight agreement, 0.21–0.40 fair agreement, 0.41–0.60 moderate agreement, 0.61–0.80 substantial agreement, and 0.81–1.00 almost perfect agreement. Statistical significance was determined using two-tailed tests with a threshold of *p* < 0.05.

## Results

### Species distribution

A total of 606 NTM clinical isolates were obtained from patients diagnosed with NTM-PD. MAC constituted the predominant subgroup (443/606, 73.1%), followed by MABC (75/606, 12.4%), *M. kansasii* (49/606, 8.1%), *M. fortuitum* (11/606, 1.8%), *M. chelonei* (6/606, 1.0%), and other rare NTM species (22/606, 3.6%). Antimicrobial susceptibility testing was performed on all isolates (Figure 1).

**Figure 1.**
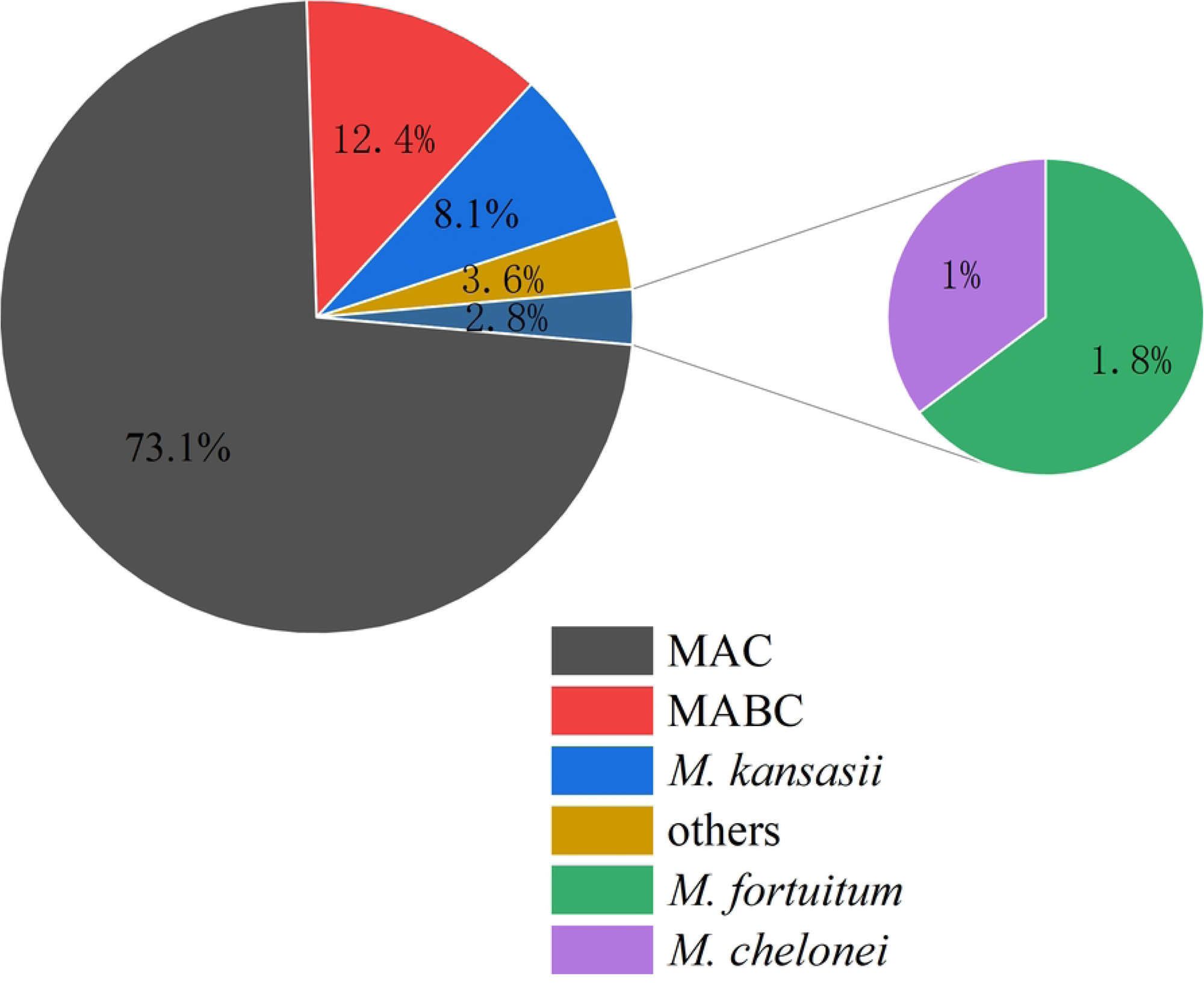
Distribution of NTM Isolates.

### Age and Gender profile in NTM-PD Patients

The cohort comprised 606 patients with NTM-PD, consisting of 332 males (54.79%) and 274 females (45.21%). The overall mean age was 58.59 ± 14.51 years (Mean ± SD). Males exhibited an older age distribution (59.8 ± 14.7 vs 57.1 ± 14.2 years, *t* = 2.31*, P* = 0.021) (Figure 2A). Among 443 MAC patients (male predominance: 52.6%, 233/443), the mean age was 61.6 ± 12.5 years. Males were significantly older than females (63.1 ± 12.7 vs. 60.0 ± 12.1 years; *t* = 2.62, *P* = 0.009) (Figure 2B). The 75 MABC patients included 27 males (36.0%) and 48 females (64.0%). The mean age was 51.5 ± 18.0 years. Although males tended to be older (55.8 ± 20.4 vs. 49.1 ± 15.9 years), this difference was not statistically significant (*t* = 1.48, *P* = 0.147) (Figure 2C). The cohort comprised 49 *M. kansasii* patients with pronounced male predominance (89.8%, 44/49). The overall mean age was 45.0 ± 12.6 years, revealing marked sex-based disparities: male patients demonstrated significantly older age distribution compared to females (46.4 ± 12.1 vs 32.6 ± 10.2 years, *t* = 2.82, *P* = 0.034) (Figure 2D). Isolates of *M. fortuitum* and other NTM species were excluded from demographic analyses owing to insufficient sample size (n <20 per subgroup).

**Figure 2.**
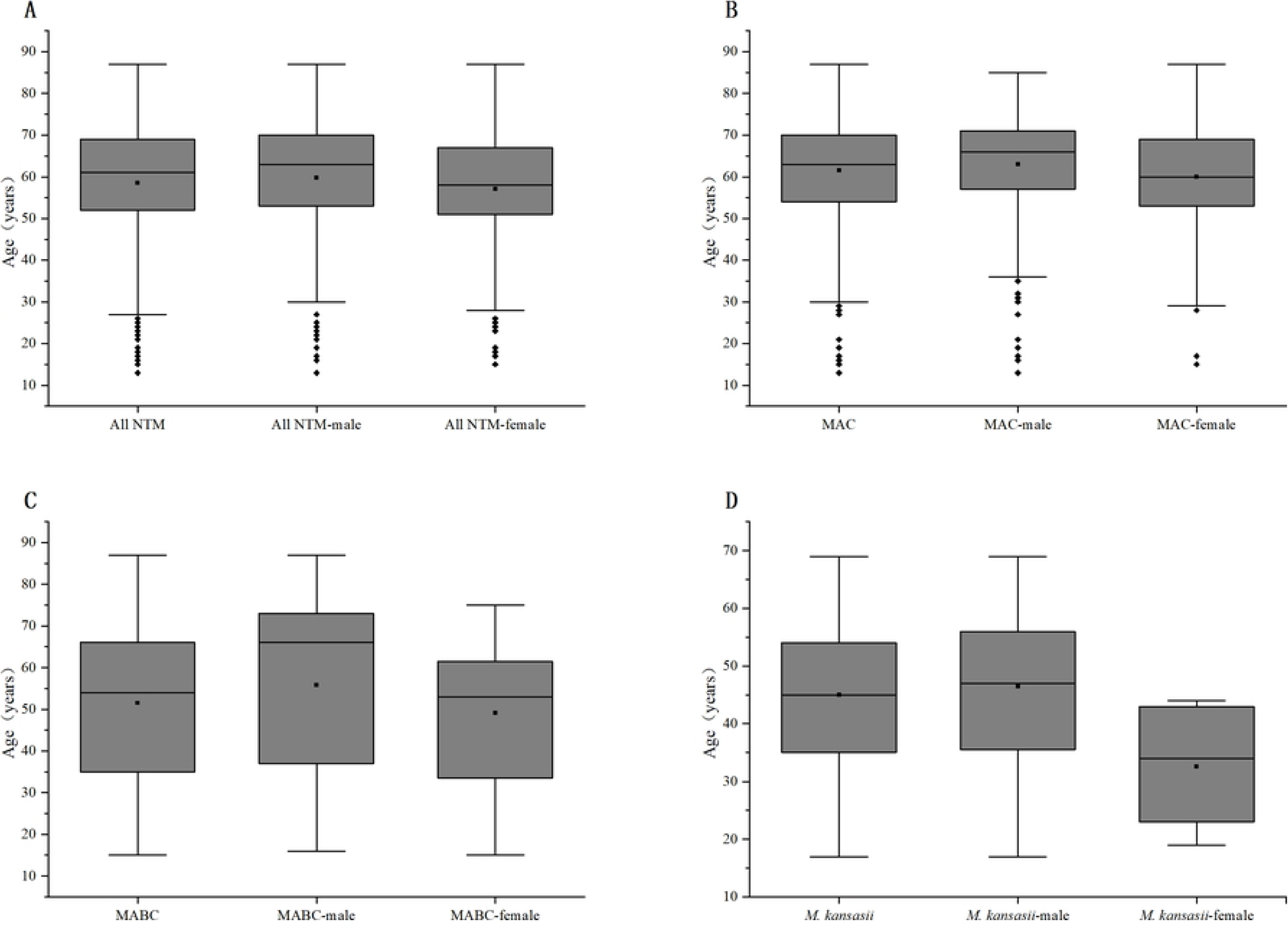
Gender-Stratified Age Distribution of NTM Isolates: Overall and Major Species. Box plots illustrate age distribution of NTM-PD patients. Panel A: Overall NTM cohort (male/female subgroups). Panels B–D: Major species (MAC, MABC, *M. kansasii*), each with gender subgroups. Box = Interquartile range (IQR, 25th–75th percentiles), central line = median, whiskers = 1.5×IQR, ■ = mean, ♦ = outliers.

### Antimicrobial susceptibility profiles of *MAC, MABC, M. kansasii and M. fortuitum* (Table 1)

This study systematically analyzed in vitro AST and resistance profile for our clinically significant mycobacterial species (MAC, MABC*, M. kansasii and M. fortuitum*) isolated from 578 clinical specimens. Linezolid demonstrated > 97% susceptibility across all four mycobacterial species. Subsequent analysis revealed that amikacin, clarithromycin, moxifloxacin, and cefoxitin exhibited notable susceptibility profiles, though marginally lower than linezolid. AST for *M. kansasii* was not conducted with imipenem, doxycycline, and minocycline, so notably excluding *M. kansasii* isolates, the antimicrobial resistance patterns for imipenem, doxycycline, and minocycline displayed pronounced resistance characteristics, which demonstrated relatively high antimicrobial resistance profiles across all four mycobacterial species. Within the MAC, imipenem and tetracyclines (including doxycycline and minocycline) demonstrate resistance rates surpassing 90%, clarithromycin, rifampicin, rifabutin, amikacin, linezolid, moxifloxacin, and cefoxitin demonstrate susceptibility rates exceeding 95%. However, azithromycin, a structurally related macrolide, exhibits markedly lower susceptibility (30.25%) and higher resistance (21.44%), presenting a striking divergence from clarithromycin, Clarithromycin demonstrated markedly superior susceptibility compared to azithromycin (98.70% versus 30.25%; *P* < 0.01). Similarly, gatifloxacin, a fluoroquinolone congener of moxifloxacin, displays a susceptibility rate of 22.35% alongside substantially elevated resistance levels compared to moxifloxacin. Ethambutol, designated as one of the three first-line therapies in the ATS guidelines for MAC pulmonary disease (MAC-PD), demonstrates low susceptibility and high resistance rates among MAC isolates, with a susceptibility rate of 38.83% classified as intermediate. Antimicrobial susceptibility testing for isoniazid was not performed in *M. kansasii* isolates. The analysis demonstrated a resistance rate of 12.25% to ethambutol, while susceptibility rates exceeding 95% were observed for both rifampicin and rifabutin. Complete susceptibility (100%) was documented for clarithromycin, amikacin, and linezolid. Fluoroquinolone agents including moxifloxacin and gatifloxacin exhibited susceptibility rates above 95%. Notably, a high resistance rate of 65.31% was observed for sulfamethoxazole. The antimicrobial susceptibility profile of MABC demonstrated complete susceptibility to linezolid and amikacin (100%), with clarithromycin, amikacin, and moxifloxacin maintaining susceptibility rates exceeding 80%. Conversely, this pathogen exhibited high resistance rates to imipenem (97.33%), doxycycline, and minocycline, all demonstrating resistance levels above 80%. Antimicrobial susceptibility testing revealed *M. fortuitum* demonstrated 100% susceptibility to amikacin, linezolid, moxifloxacin, and cefoxitin. Clarithromycin showed reduced efficacy with 81.80% susceptibility, the resistance rates of imipenem and doxycycline are as high as over 90%.

**Table 1.** Antimicrobial Susceptibility Profiles of Different Drugs Against Mycobacterial Isolates.

| Drugs/<br>Isolates | MAC |  | MABC |  | *M. Kansasii* |  | *M. fortuitum* |  |
| --- | --- | --- | --- | --- | --- | --- | --- | --- |
|  | n | % | n | % | n | % | n | % |
|  | (S/I/R) | (S/I/R) | (S/I/R) | (S/I/R) | (S/I/R) | (S/I/R) | (S/I/R) | (S/I/R) |
| RIF | 441/1/1 | 99.55/0.23/0.23 |  |  | 47/0/2 | 95.92/0/4.08 |  |  |
| CLA | 438/0/5 | 98.87/0/1.13 | 66/8/1 | 88/10.67/1.33 | 49/0/0 | 100/0/0 | 9/2/0 | 81.80/18.20/0 |
| RFB | 437/0/6 | 98.65/0/1.36 |  |  | 48/0/1 | 97.96/0/2.04 |  |  |
| AMK | 435/6/2 | 98.19/1.35/0.45 | 75/0/0 | 100/0/0 | 49/0/0 | 100/0/0 | 10/0/1 | 90.90/0/9.10 |
| LZD | 430/11/2 | 97.07/2.48/0.45 | 75/0/0 | 100/0/0 | 49/0/0 | 100/0/0 | 11/0/0 | 100/0/0 |
| MXF | 428/15/0 | 96.61/3.39/0 | 61/9/5 | 81.33/12/6.67 | 48/0/1 | 97.96/0/2.04 | 10/0/1 | 90.90/0/9.10 |
| FOX | 424/10/9 | 95.71/2.26/2.03 | 61/9/1 | 86.49/12.16/1.35 |  |  | 11/0/0 | 100/0/0 |
| TOB | 179/243/21 | 40.41/54.85/4.74 | 3/17/55 | 4/22.67/73.33 |  |  | 1/2/8 | 9.10/18.20/72.70 |
| AZM | 134/214/95 | 30.25/48.31/21.44 | 29/17/10 | 51.79/30.36/17.86 |  |  |  |  |
| GAT | 99/167/177 | 22.35/37.70/39.95 |  |  | 47/0/2 | 95.92/0/4.08 |  |  |
| EMB | 77/172/194 | 17.38/38.83/43.79 |  |  | 41/2/6 | 83.67/4.08/12.25 |  |  |
| SXT | 77/13/353 | 17.38/2.93/79.68 | 17/5/53 | 22.67/6.67/70.67 | 15/2/32 | 30.61/4.08/65.31 | 4/0/7 | 36.36/0/63.64 |
| IMP | 40/1/402 | 9.03/0.23/90.74 | 1/1/73 | 1.33/1.33/97.33 |  |  | 1/0/10 | 9.10/0/90.90 |
| DOX | 9/9/425 | 2.03/2.03/95.94 | 7/3/64 | 9.46/4.05/86.49 |  |  | 1/0/10 | 9.10/0/90.90 |
| MIN | 7/8/428 | 1.58/1.81/96.61 | 9/2/45 | 16.07/3.57/80.36 |  |  |  |  |

n (S/I/R): Number of isolates categorized as Susceptible (S), Intermediate (I), and Resistant (R).
%(S/I/R): Proportion of isolates with Susceptible/Intermediate/Resistant phenotypes.
Drug abbreviations: RIF (rifampin), RFB (rifabutin), CLA (clarithromycin), AMK (amikacin), LZD (linezolid), MXF
(moxifloxacin), SXT (trimethoprim-sulfamethoxazole), FOX (cefoxitin), GAT (gatifloxacin), TOB (tobramycin),
AZM (azithromycin).

*M. chelonei* was isolated from six clinical specimens, reflecting a limited sample size. AST (susceptible/intermediate/resistant) demonstrated uniform susceptibility (6/0/0) to amikacin, moxifloxacin, clarithromycin, and linezolid. Cefoxitin demonstrated susceptibility in 5 isolates (5/1/0), with sulfamethoxazole susceptibility observed in 5 cases (5/0/1). Minocycline susceptibility was recorded as 4/0/2. Resistance profiles displayed the following distributions: tobramycin (1/2/3), azithromycin (1/1/4), doxycycline (1/1/4), and imipenem (0/0/6).

### Drug combination susceptibility in MAC and MABC isolates

Based on AST results, this investigation systematically assessed 3–6 agent regimens against MAC and MABC. Quantitative analysis of synergistic efficacy profiles was performed, followed by hierarchical stratification of combinations demonstrating superior bactericidal potential.

Analysis of triple regimens against MAC isolates identified the top 10 most sensitive regimens, predominantly consisting of rifamycin-clarithromycin paired with moxifloxacin, amikacin, linezolid, or cefoxitin, each antimicrobial agent demonstrated susceptibility rates surpassing 95% in MAC susceptibility testing. Susceptibility rates among the leading regimens ranged from 92.6% (410/443) to 96.6% (428/443), with comparative analysis demonstrating no statistically significant variation (*p* > 0.05), These findings suggest no significant differences in susceptibility rates observed among the top ten triple regimens. The optimized Quadruple drug regimens (rifamycins + clarithromycin combined with linezolid, cefoxitin and moxifloxacin) demonstrated susceptibility rates ranging from 91.4% (405/443) to 93.7% (415/443), with comparative analysis revealing no significant intergroup variation (*P* > 0.05). The optimized quintuple therapeutic regimens demonstrated maximal antimicrobial activity through combinations of rifamycin derivatives, clarithromycin, linezolid, cefoxitin, and moxifloxacin. There is no statistical difference between the highest and lowest susceptibility of the five-drug combination (*P* > 0.05), which indicates that there is no statistical significance among the top ten five-drug combinations with the highest susceptibility. The two leading six-drug combination regimens in terms of susceptibility are those that include rifamycins combined with clarithromycin, amikacin, linezolid, moxifloxacin, and cefoxitin. These regimens exhibit susceptibility rates of 86.91% (385/443) and 86.00% (381/443), respectively. No statistically significant difference was observed between the two regimens. The remaining six drug combinations with higher susceptibility rates are also based on rifamycins, clarithromycin, linezolid, cefoxitin, moxifloxacin, and amikacin in combination with tobramycin, azithromycin, gatifloxacin, and sulfamethoxazole (Figure 3A–D). Analysis of MABC isolates revealed susceptibility patterns mirroring MAC with optimized 3–6 drug regimens incorporating linezolid, clarithromycin, moxifloxacin, cefoxitin, and amikacin (individual susceptibility > 80%), The optimized therapeutic regimen incorporating the aforementioned antimicrobial agents, when augmented with azithromycin and sulfamethoxazole, demonstrated preferred susceptibility profiles (Figure 4A–D).

**Figure 3.**
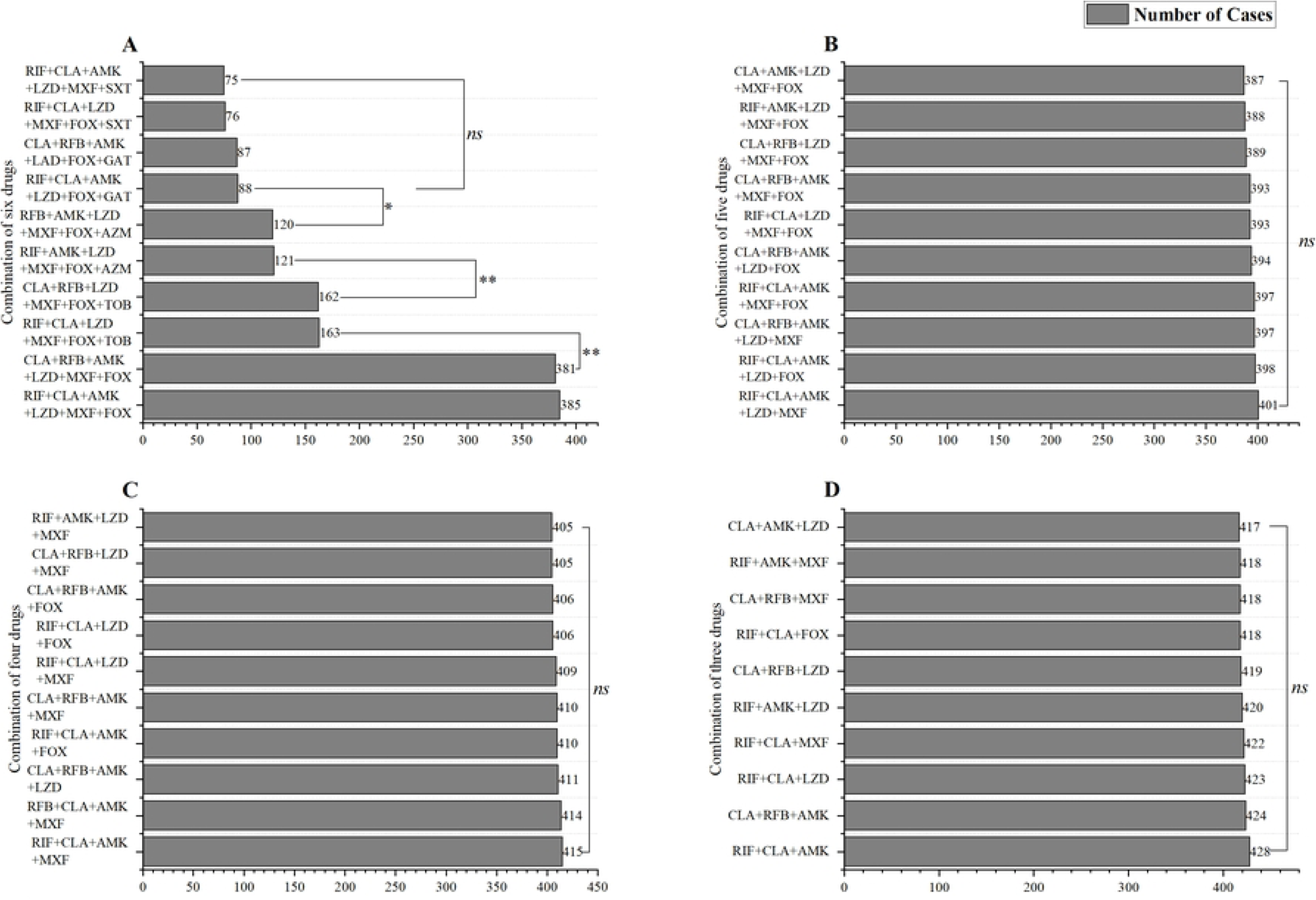
Multi-Drug Combinations Susceptibility Rankings in MAC-PD Patients: 3–6 Drug Regimens. Bar charts illustrate the distribution of MAC-PD patients exhibiting full susceptibility to multi-drug combinations, categorized by regimens containing 6 (A), 5 (B), 4 (C), and 3 drugs (D). Chi-square tests determined statistical significance: * *P* < 0.05, ** *P* < 0.01, ns = no statistical significance.

**Figure 4.**
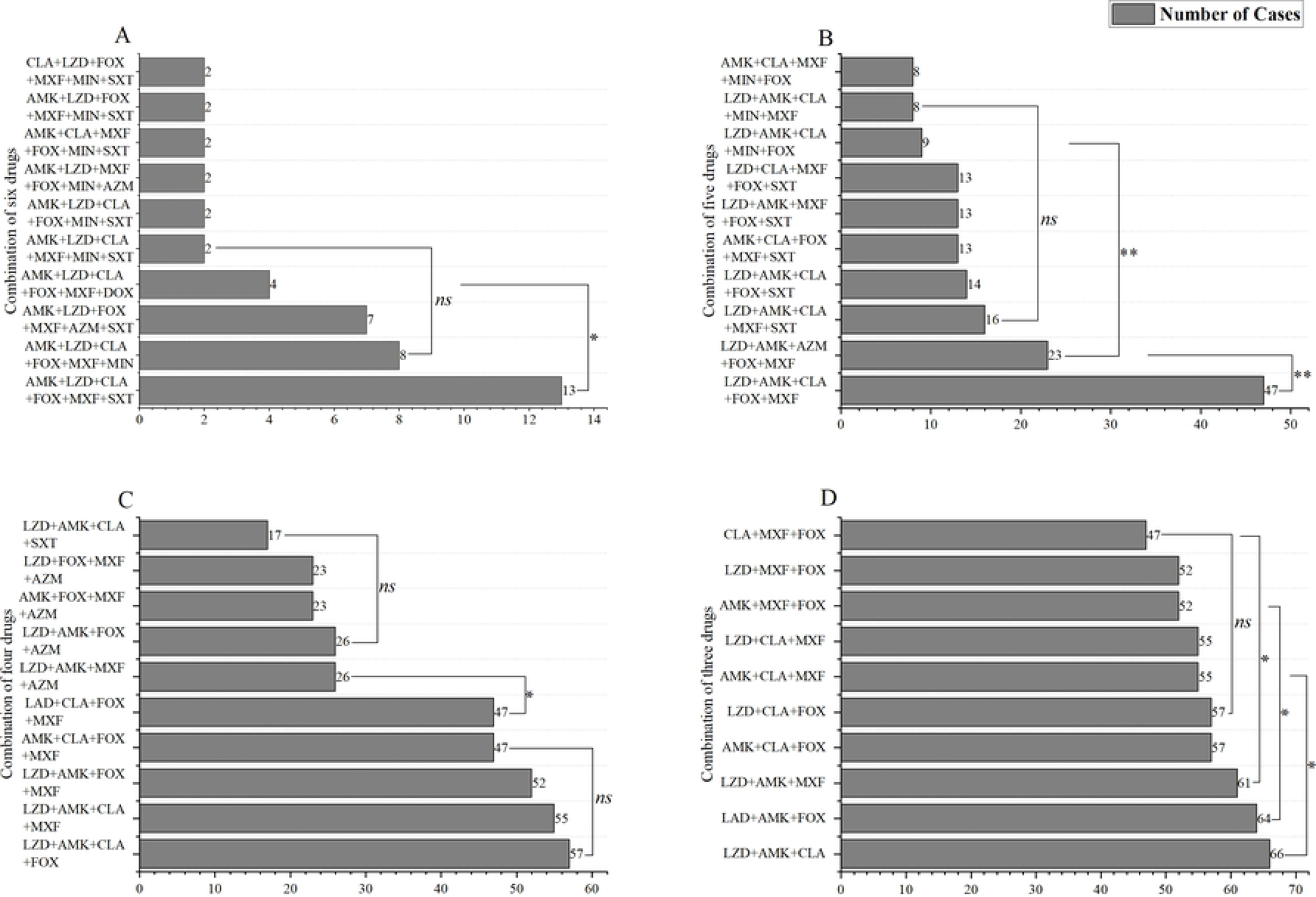
Multi-Drug Combinations Susceptibility Rankings in MABC-PD Patients: 3–6 Drug Regimens. Bar charts illustrate the distribution of MABC-PD patients exhibiting full susceptibility to multi-drug combinations, categorized by regimens containing 6 (A), 5 (B), 4 (C), and 3 drugs (D). Statistical analysis followed the same methodology as in Figure 3. Significant differences were observed between non-adjacent groups, while intermediate groups showed no significant differences among themselves or with non-adjacent groups. Due to layout constraints, differences between groups were not annotated in the graph for logical clarity. Abbreviations follow the definitions in Figure 3, including a new drug (min = minocycline).

### Correlation among congeneric therapeutic agents

Comparative analysis of MAC isolates demonstrated differential antimicrobial susceptibility profiles (Table 2). Clarithromycin exhibited superior susceptibility rates compared to azithromycin, with the latter showing elevated intermediate frequencies, no intermediate susceptibility was observed for clarithromycin. Statistical measures revealed minimal association between these agents (Cramér’s V = 0.155, κ = 0.011), indicating weak correlation and poor concordance. Both rifampicin and rifabutin maintained high susceptibility rates, demonstrating moderate inter-drug correlation yet discordant susceptibility patterns. Amikacin showed significantly greater susceptibility than tobramycin, with negligible correlation. Moxifloxacin demonstrated enhanced susceptibility relative to gatifloxacin, exhibiting moderate association but no significant consistency, this may be attributed to moxifloxacin resistance rate being 0 and its intermediate rate being exceptionally low.

**Table 2.**
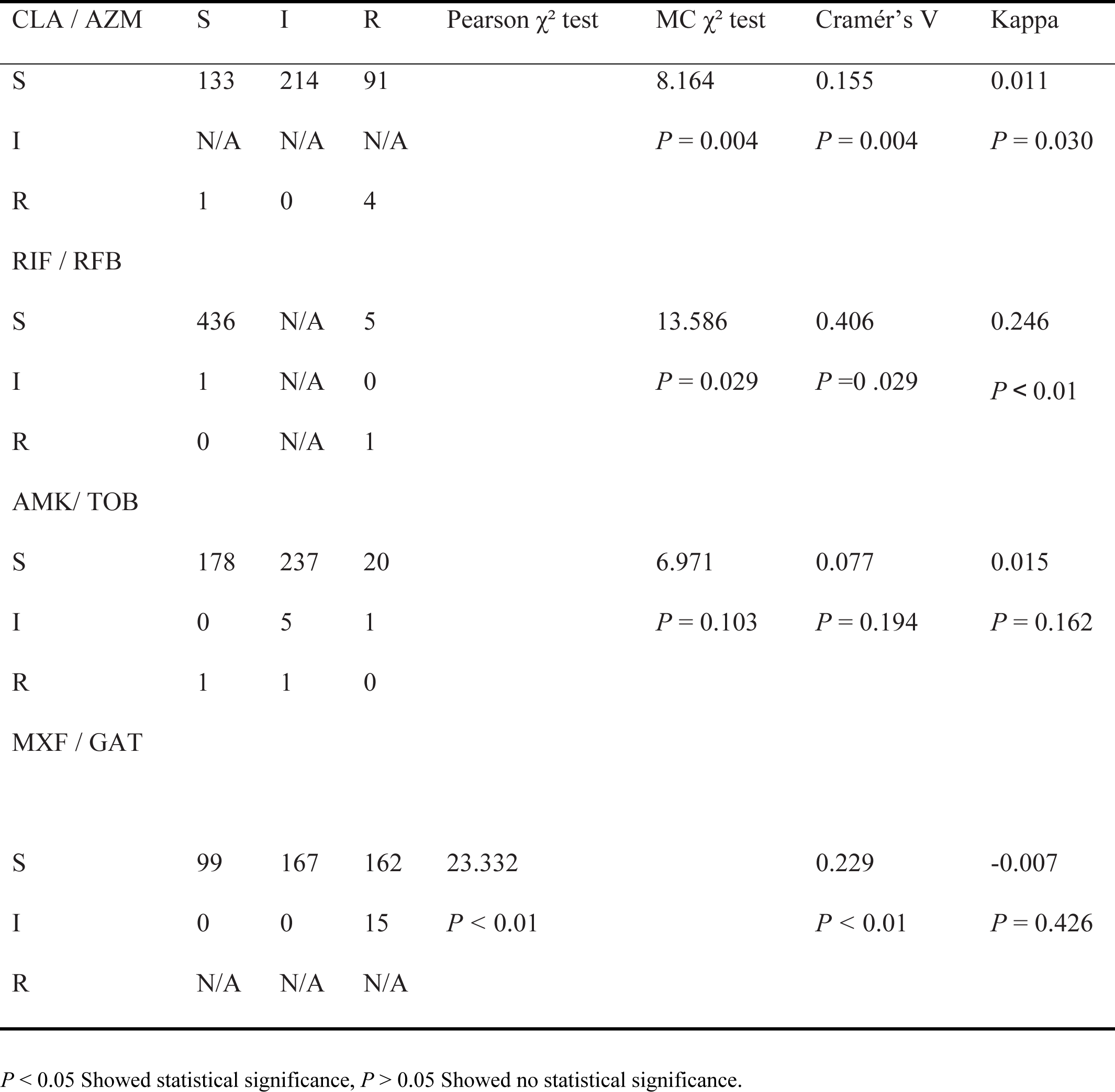
Association Analysis of Antimicrobial Susceptibility Patterns Among Congeneric Therapeutic Agents in MAC Isolates.

Among MABC isolates (Table 3), clarithromycin demonstrates higher susceptibility rates than azithromycin, with moderate correlation but poor agreement between these agents. Minocycline and doxycycline exhibit elevated resistance rates, showing moderate correlation and substantial consistency. The 100% susceptibility rate of amikacin precludes a conclusive assessment of its correlation with tobramycin.

**Table 3.** Association Analysis of Antimicrobial Susceptibility Patterns Among Congeneric Therapeutic Agents in MABC isolates.

| CLA/AZM | S | I | R | Pearson $\chi^2$ test | MC $\chi^2$ test | Cramér's V | Kappa |
| --- | --- | --- | --- | --- | --- | --- | --- |
| S | 29 | 16 | 6 |  | 10.816 | 0.516 | 0.074 |
|  |  |  |  |  | *P* = 0.001 | *P* = 0.001 | *P* < 0.01 |
| I | 0 | 1 | 4 |  |  |  |  |
| R | N/A | N/A | N/A |  |  |  |  |
| MIN/DOX |  |  |  |  |  |  |  |
| S | 9 | 3 | 3 |  | 36.362 | 0.580 | 0.622 |
|  |  |  |  |  | *P* < 0.01 | *P* < 0.01 | *P* < 0.01 |
| I | 0 | 0 | 2 |  |  |  |  |
| R | 1 | 0 | 43 |  |  |  |  |
| AMK/TOB |  |  |  |  |  |  |  |
| S | 3 | 17 | 55 |  |  |  |  |
| R | N/A | N/A | N/A |  |  |  |  |

*P* < 0.05 Showed statistical significance, *P* > 0.05 Showed no statistical significance.

### Antimicrobial susceptibility comparison between slow growing mycobacteria (SGM) and rapid growing mycobacteria (RGM)

The NTM strains can be categorized into two distinct classes, slow-growing and fast-growing mycobacteria, based on their distinct growth rates. The SGM in this study predominantly comprised MAC and *M. kansasii*, while the RGM consisted of MABC, *M. fortuitum*, and *M. chelonei*. Antimicrobial susceptibility testing was performed on a panel of 16 agents against MAC isolates. Testing for azithromycin, tetracyclines, imipenem, cefoxitin, and tobramycin was not performed with M. kansasii isolates. Clarithromycin, rifamycins, moxifloxacin, and linezolid demonstrate favorable susceptibility profiles against both MAC and *M. kansasii*. Nevertheless, gatifloxacin and ethambutol show higher susceptibility and lower resistance in *M. kansasii*, but present relatively low susceptibility in MAC. Notably, sulfamethoxazole displays substantial resistance in both mycobacterial species, with particularly elevated resistance rates observed in MAC isolates. Antimicrobial susceptibility testing revealed differential patterns among mycobacterial species: MABC and *M. chelonei* demonstrated susceptibility to 11 therapeutic agents, whereas *M. fortuitum* retained susceptibility to 9 antimicrobials and the tests for azithromycin and minocycline could not be conducted. Linezolid, amikacin, clarithromycin, cefoxitin, and moxifloxacin demonstrated high susceptibility profiles in all RGM isolates, while limited to small sample sizes, *M. chelonei* isolates showed favorable susceptibility to sulfamethoxazole and minocycline with minimal resistance, notably, MABC and *M. fortuitum* exhibited elevated resistance profiles to tobramycin and imipenem. Tobramycin and imipenem demonstrated elevated resistance rates among all three RGM species. Notably, doxycycline resistance was particularly pronounced in MABC and *M. chelonei* isolates.

## Discussion

The historical misdiagnosis of tuberculosis, due to inadequate isolation methods and outdated clinical paradigms, has had severe consequences. Patients endured ineffective antituberculosis chemotherapy, leading to disease progression, development of acquired resistance to drugs like rifamycins and ethambutol, and the emergence of refractory NTM-PD, complicating long-term management and prognosis. International NTM treatment guidelines emphasize the importance of AST, specifying susceptibility testing for key drugs against major NTM pathogens, such as macrolides for MAC and MABC, and rifamycins for *M. kansasii*[3,6]. However, the clinical efficacy of numerous drugs, including rifamycins (for MAC), linezolid, and quinolones, remains incompletely validated *in vivo*. Therefore, it is essential to enhance NTM susceptibility testing. This enhancement serves not only to provide a foundation for developing clinical treatment plans but also to supply crucial data for further clinical trials. Additionally, regional-level susceptibility analysis can support empirical treatment strategies for NTM diseases within the region.

The global epidemiology of NTM infections demonstrates significant geographical heterogeneity, with emerging trends in both microbial isolation rates and clinical disease incidence[11]. Surveillance data reveal distinct regional patterns in NTM-PD prevalence. North American studies document an ascending trajectory of NTM-PD cases, particularly in southeastern and southwestern United States regions characterized by warm, humid climates. Canadian epidemiological monitoring indicates a notable surge in MAC pulmonary disease (MAC-PD) incidence within Ontario province. European epidemiological profiles exhibit substantial intercountry variability in NTM isolation frequencies and disease burden, with MAC and MABC predominating as principal pathogens. The Central/North American and South American regions demonstrate fragmented surveillance, lacking comprehensive epidemiological data. African NTM-PD reporting remains sparse, while East Asian nations including Japan and South Korea report progressive increases in disease incidence, predominantly associated with MAC and MABC infections. The 2010 report of the fifth national tuberculosis epidemic survey in China shows that the isolation rate of NTM increased significantly from 4.9% in 1990 to 22.9% in 2010. A 2021 nationwide multicenter analysis of tuberculosis referral centers in China documented NTM-PD accounting for 6.8% of culture-confirmed pulmonary mycobacteriosis cases[12]. Another study conducted in China demonstrated that geospatial analysis revealed significant latitudinal gradients in both mycobacterial isolation rates and NTM-PD incidence across mainland China, with differential species distribution: SGM demonstrated northern latitudinal predominance, contrasting with RGM showing epidemiological dominance in southern provinces[13]. This study identified MAC, MABC*, M. kansasii, M. fortuitum*, and *M. chelonei* as predominant NTM species isolated from pulmonary specimens at our institution. As the designated tuberculosis referral center for Shandong Province, China, our hospital receives patients from diverse geographical regions across this 157,900 km² administrative area and neighboring jurisdictions. Situated in eastern China (34°25’– 38°43’N), Shandong’s climate exhibits spatial variability predominantly characterized by temperate continental monsoon conditions with distinct seasonal patterns, while coastal regions demonstrate modified oceanic influences featuring elevated humidity levels. These environmental parameters may influence the ecological distribution of NTM species within the province’s microbial reservoirs. NTM-PD demonstrates distinct epidemiological patterns across geographical regions. Globally, the condition predominantly affects individuals aged over 50 years, with East Asian populations exhibiting elevated patient ages compared to other regions. Japanese and Korean cohorts demonstrate mean onset at 60–70 years with notable female predominance (65%–88%) in MAC-PD, contrasting with balanced gender distributions observed in Western populations. MABC lung disease (MABC-PD) shows marginal female predominance in East Asia, while maintaining gender parity in European and American studies. Notably, *M. kansasii* lung diseases demonstrate male predominance in European populations. This study reveals that among all cases of NTM-PD, the proportion of male patients is slightly higher than that of female patients, and the age of onset for males is generally higher than for females. This finding contrasts with data from Japan and South Korea, where the average age of onset falls within the 60–70 years range. Given that the economic development level of Shandong Province in China lags behind that of developed countries such as Japan and South Korea, it is hypothesized that the incidence of NTM-PD may be associated with the level of economic development. In the case of MAC-PD, the majority of patients are elderly males, whereas pulmonary disease caused by MABC predominantly affects elderly females, with a lower average age of onset compared to MAC-PD. Additionally, the number of male patients with *M. kansasii* infection significantly exceeds that of female patients, and the average age of onset is relatively younger in both genders, with females exhibiting an earlier onset age than males. Epidemiologic evidence indicates geographic variations in the age and sex distribution of NTM-PD onset, with potential associations to ethnic differences. Comparative analyses reveal demographic variations in age and sex distribution among NTM species associated with pulmonary disease.

This study performed statistical analysis of antimicrobial susceptibility profiles among patients with NTM-PD infected by MAC, MABC*, M. kansasii,* and other infrequent mycobacterial species. Susceptibility testing evaluated therapeutic agents across major antimicrobial classes such as macrolides, fluoroquinolones, quinolones, and aminoglycosides etc. The management of NTM-PD necessitates multidrug regimens due to high treatment failure rates, with disseminated cases complicated by immunodeficiency often requiring 5–6-agent combinations. However, clinical implementation faces substantial barriers including intrinsic or acquired resistance to first-line agents, drug allergies, and dose-limiting toxicities from prolonged polypharmacy. As next-generation sequencing technologies become increasingly utilized in diagnostic workflows, many patients receive confirmed NTM-PD diagnoses before antimicrobial susceptibility reports become available. To address this therapeutic dilemma, we systematically assessed the clinical efficacy of 3–6 drug combinations through species-specific sensitivity profiling and therapeutic hierarchy establishment. These findings provide an evidence-based framework for optimizing antimicrobial selection in our regional NTM-PD population, enabling rational empirical therapy when susceptibility data are pending while minimizing adverse outcomes associated with inappropriate regimen choices.

The MAC represents the predominant etiological agent of NTM-PD globally and maintains its epidemiological dominance in China[1,12], though recent epidemiological investigations position it as secondary to the MABC in certain regional studies[14]. This complex primarily comprises *Mycobacterium intracellulare, Mycobacterium avium, Mycobacterium chimaera, and Mycobacterium paracellulare.* The development of antimicrobial resistance in MAC involves multifaceted mechanisms. A limited-scale Thai investigation (n=38) demonstrated resistance rates to amikacin, clarithromycin, linezolid, and moxifloxacin of 36.8%, 44.7%, 78.9%, and 81.6%, respectively. Resistant isolates harbored mutations in the rrs (amikacin resistance), *rrl* (clarithromycin resistance), *rplC* (linezolid resistance), and *gyrB* (moxifloxacin resistance) genes[15]. Within the MAC, resistance potentially arises through efflux pump overexpression, drug target modification, and altered cell wall permeability[16]. Certain antimicrobial agents, including macrolides and aminoglycosides, have demonstrated efficacy against MAC. Clinical studies indicate that clarithromycin and amikacin exhibit favorable susceptibility profiles in AST for NTM-PD management[17,18]. However, no statistically significant correlation has been established between in vitro susceptibility results and therapeutic outcomes for linezolid or moxifloxacin in clinical applications. National surveillance data from China demonstrated clarithromycin susceptibility in 95.37% of MAC isolates (4.63% resistance), while amikacin exhibited 71.30% susceptibility (10.19% resistance)[14]. Both linezolid and moxifloxacin showed limited effectiveness with susceptibility rates below 20%. Israeli epidemiological investigations revealed ≥ 97% susceptibility for clarithromycin and amikacin against MAC, contrasting with 25% moxifloxacin susceptibility and 3% for linezolid[19]. US multicenter studies[8] documented clarithromycin susceptibility ranging 94–100%, amikacin 64– 91%, moxifloxacin 4–41%, and linezolid 2–31% across different MAC strains. Drug susceptibility analysis in this study demonstrated high susceptibility rates (> 96%) to MAC for rifampicin, clarithromycin, amikacin, moxifloxacin, rifabutin, linezolid, and cefoxitin among clinical isolates; ethambutol exhibited markedly reduced susceptibility (approximately 17%), indicating predominant resistance characteristics. Emerging evidence indicates that macrolide resistance correlates with therapeutic failure in MAC-PD, while preserved macrolide susceptibility enhances treatment efficacy for NTM-PD[20,21]. Clinical data demonstrate that amikacin adjunctive therapy significantly improves sputum culture conversion rates in MAC-PD cases, particularly benefiting patients with unsuccessful initial therapeutic regimens[22]. Current guidelines recommend amikacin-containing combination therapies as a validated approach for managing refractory MAC-PD[6]. The ATS and British Thoracic Society guidelines mandate a triple antimicrobial regimen comprising a macrolide and ethambutol as foundational agents, supplemented by either amikacin, rifampicin, or a fluoroquinolone. These agents should be selected in accordance with antimicrobial susceptibility testing results for macrolides and amikacin[3,6]. In vitro susceptibility testing in this study demonstrated high activity for rifampicin, clarithromycin, amikacin, and moxifloxacin, whereas ethambutol exhibited markedly reduced susceptibility with elevated resistance rates. Empirical regimens guided by these susceptibility profiles should avoid routine inclusion of ethambutol in three-drug combinations. Current evidence indicates no correlation between rifampicin/ethambutol susceptibility patterns and treatment failure or relapse in MAC pulmonary disease. Notably, ATS guidelines recommend maintaining ethambutol in therapeutic regimens due to its potential role in preventing macrolide resistance development. The current investigation demonstrated that therapeutic regimens incorporating rifamycins (rifampin/rifabutin) combined with macrolides (clarithromycin/azithromycin), amikacin, linezolid, and fluoroquinolones (moxifloxacin/ciprofloxacin) as foundational agents, when supplemented with additional antimicrobials to form 3–6 drug combinations, demonstrated optimal susceptibility profiles. This antimicrobial efficacy directly correlated with baseline susceptibility patterns of individual agents against MAC clinical isolates, indicating an absence of cross-resistance or antagonistic interactions among pharmacologically distinct compound classes. While no statistically significant differences in sensitivity were observed between some drug combinations, the potential adverse effects arising from their combined use, along with the possibility of allergic reactions, highlight the critical need for comprehensive sensitivity data on drug combinations. Such information can offer valuable support for empirical treatment of MAC-PD and inform the design of future clinical trials.

MABC-PD is the second most prevalent nontuberculous mycobacterial pulmonary disease in China, surpassed only by MAC-PD. This epidemiological trend is consistent across North America, Europe, and other regions of Asia, where MABC-PD also ranks second in prevalence after MAC-PD[1]. According to the current ATS clinical guidelines[3], a macrolide-based treatment regimen is recommended for managing MABC-PD, which must be combined with intravenous antimicrobial agents (such as imipenem, cefoxitin, or amikacin) or oral antibiotics sensitive to antimicrobial susceptibility testing. The guidelines further stress the importance of conducting susceptibility testing for both macrolides and aminoglycosides, with therapeutic decisions being guided by antimicrobial sensitivity profiles to optimize the composition of multidrug regimens. A recent Israeli investigation evaluated antimicrobial susceptibility patterns of MABC strains in Israel and selected Middle Eastern regions, among tested agents, amikacin showed superior efficacy with 98% susceptibility, contrasting sharply with linezolid and clarithromycin, which demonstrated suboptimal activity (<50% susceptibility). Notably, imipenem exhibited limited effectiveness at 11% susceptibility, 77% intermediary rate, while resistance patterns revealed 18% full resistance[19]. Parallel data from the United Kingdom revealed divergent susceptibility trends, Tigecycline emerged as the most potent agent with 99% susceptibility, followed by clarithromycin at 94%. Conversely, ciprofloxacin, imipenem, linezolid, and cefoxitin all demonstrated minimal clinical utility with susceptibility rates below 5%[23]. Chinese surveillance data from an extensive multicenter investigation revealed distinct antimicrobial susceptibility patterns among MABC isolates, amikacin demonstrated superior activity with 94.74% susceptibility, followed by clarithromycin at 85.09%. In contrast, linezolid exhibited limited efficacy (32.46% susceptibility). Notably elevated resistance rates were observed for quinolones, tobramycin, and imipenem, with susceptibility levels falling below clinically acceptable thresholds[14]. In vitro antimicrobial susceptibility analysis of this study revealed clarithromycin maintained 88.00% efficacy within macrolides, while azithromycin exhibited reduced sensitivity (51.79% susceptibility) with emerging resistance (17.00%). Notably, 97.33% of isolates demonstrated resistance to imipenem. The antimicrobial sensitivity profile ranked as follows: linezolid and amikacin showed complete susceptibility (100%), followed by clarithromycin (88.00%), cefoxitin (86.49%), moxifloxacin (81.33%), and azithromycin (51.79%). Mycobacterium abscessus complex comprises three distinct subspecies: *M. abscessus subsp. abscessus* (MAB), *M. abscessus subsp. massiliense* (MMA), and *M. abscessus subsp. bolletii* (MBO). Critical to macrolide resistance mechanisms, the inducible *erm*(41) gene mediates mutational alterations in the 23S rRNA gene upon clarithromycin exposure. Genetic characterization reveals MABC specifically harbors a functional *erm(41)* allele, correlating with its capacity to develop acquired macrolide resistance through ribosomal target modification. Antimicrobial susceptibility patterns demonstrate temporal variability in resistance development. Initial susceptibility testing may indicate sensitivity within 3-day incubation periods, while extended incubation (>14 days) reveals emergent clarithromycin resistance in *erm(41)*-positive strains. Comparative genomic analysis identifies intact *erm(41)* sequences in both MAB and MBO subspecies, enabling inducible macrolide resistance through ribosomal methylation[24]. Conversely, MMA exhibits truncated *erm(41)* sequences that preclude functional resistance induction. This differential genetic architecture explains observed variations in clinical treatment outcomes, emphasizing the necessity for extended antimicrobial susceptibility testing protocols when managing infections caused by *erm*(41)-positive subspecies. Distinct MABC subspecies exhibit variable resistance profiles. MAB (76.7%) and MBO (100%) subspecies display high clarithromycin resistance, primarily mediated by *erm(41)* activation with rare *rrl* mutations. Conversely, MMA exhibits lower clarithromycin resistance (10.6%), also attributable to *erm(41)*. Amikacin resistance across all three subspecies remains below 7%, uniformly associated with *rrs* mutations[25]. MABC demonstrates intrinsic resistance to primary antitubercular medications while exhibiting broad-spectrum antimicrobial resistance encompassing β-lactams, aminoglycosides, and tetracyclines[26]. The management of MABC pulmonary infections presents considerable therapeutic obstacles, with documented treatment success rates remaining below 50% in clinical settings. Clinical investigations demonstrate significant concordance between clarithromycin and azithromycin in vitro susceptibility profiles and corresponding therapeutic outcomes[3], with amikacin exhibiting analogous clinical correlations[23]. In contrast, most antimicrobial agents display limited predictive validity of susceptibility testing for clinical response. Consequently, current ATS NTM Guidelines prioritize therapeutic agent selection based on clarithromycin and amikacin susceptibility patterns. Macrolide therapeutic outcomes in MABC-PD exhibit subspecies-dependent variability, despite these agents serving as primary antimicrobial therapy. Current taxonomic delineation lacks critical strain-specific characterization, highlighting the necessity for refined genomic subtyping protocols integrated with *erm*(41) gene analysis to optimize clinical management strategies. Current therapeutic protocols for MABC-PD endorse multidrug regimens comprising 3–4 antimicrobial agents, with intensive-phase regimens occasionally extending to five agents. While the 2020 ATS guidelines emphasize this combinatorial approach, the 2017 British Thoracic Society framework advocates expanded susceptibility profiling for some therapeutic classes (macrolides, aminoglycosides, β-lactams, carbapenems, fluoroquinolones, oxazolidinones, and riminophenazines) to inform therapeutic strategies, though strictly as advisory parameters. The investigation established that the susceptibility profiles of amikacin, linezolid, clarithromycin, fluoroquinolones, and cefotiam demonstrated efficacy rates exceeding the 80% threshold. Multidrug protocols incorporating 3–6 agents from these pharmacological classes demonstrated maximal antimicrobial susceptibility profiles. Consequently, these antimicrobial agents represent preferred therapeutic options when formulating regimens guided by in vitro susceptibility testing data.

*M. kansasii* pulmonary disease demonstrates a comparatively low epidemiological prevalence. Current therapeutic strategies parallel those employed in pulmonary tuberculosis management. The ATS 2020 guidelines advocate a rifampin-based regimen combined with ethambutol and either isoniazid or clarithromycin for antimycobacterial therapy[3]. Notably, the guidelines explicitly advise against aminoglycosides and fluoroquinolones as primary therapeutic agents. Fluoroquinolone administration is strictly reserved for cases demonstrating rifampin or isoniazid intolerance, or when clinical contraindications preclude their utilization. The exclusion of aminoglycosides from recommended protocols stems from the established efficacy of oral triple-drug therapy, thereby obviating the necessity for amikacin administration which would unnecessarily elevate the risk of adverse effects while imposing undue clinical burden. Molecular studies indicate that M. kansasii resistance frequently correlates with specific genetic mutations: *rpoB* mutations (notably *S454L*) confer high-level rifampin resistance[27], *A2266C* in *rrl* associates with clarithromycin resistance, and *A128G* in *rpsL* links to streptomycin resistance[28]. Intrinsic resistance to ethambutol and isoniazid occurs in some strains[27], while active drug efflux mechanisms reduce intracellular antibiotic concentrations, further potentiating resistance[28]. Chinese data demonstrated clarithromycin, rifabutin, amikacin, linezolid, and moxifloxacin susceptibility rates exceeding 96%, with rifampin showing 93.55% efficacy. Sulfamethoxazole exhibited markedly reduced sensitivity at 48.39%, while ciprofloxacin and doxycycline displayed elevated resistance profiles[14]. Israeli investigations revealed comparable antimicrobial susceptibility patterns to Chinese findings, though with distinct exceptions: trimethoprim-sulfamethoxazole sensitivity reached 88% and ciprofloxacin susceptibility improved to 74% in Israeli isolates[19]. United States surveillance data indicated clarithromycin and rifabutin maintained high susceptibility rates (98%), followed by amikacin at 90% efficacy. Reduced susceptibility emerged for rifampin, moxifloxacin, and ciprofloxacin (≤ 80%), while trimethoprim-sulfamethoxazole mirrored Chinese resistance patterns at 48% susceptibility. Ciprofloxacin, doxycycline, and minocycline consistently demonstrated elevated resistance rates across all studied populations[8]. Cross-regional antimicrobial susceptibility analyses demonstrate consistent resistance trends despite geographical variations in specific resistance profiles. The antimicrobial susceptibility profile of *M. kansasii* observed in this investigation aligned with findings from three prior studies. Clarithromycin, amikacin, linezolid, rifabutin, moxifloxacin, rifampin, and gatifloxacin each demonstrated susceptibility rates exceeding 95%, whereas ethambutol exhibited reduced efficacy at 83.7%. Notably, sulfamethoxazole displayed markedly diminished susceptibility compared to other tested agents. Given the high susceptibility profile demonstrated by *M. kansasii* isolates in antimicrobial testing, this investigation did not pursue a comparative assessment of combination drug therapeutic efficacy. The ATS clinical guidelines recommend a triple-drug regimen comprising isoniazid, rifampicin, and ethambutol administered over 9 to 18 months for pulmonary disease caused by *M. kansasii,* demonstrating cure rates of 80-100% with favorable long-term outcomes. Two limited retrospective cohort investigations evaluating clarithromycin substitution for isoniazid in comparable regimens reported analogous therapeutic efficacy, with sustained mycobacterial eradication achieved in 80–100% of cases across both study populations. Consequently, the guidelines advocate a therapeutic regimen comprising rifampin, isoniazid, ethambutol, and clarithromycin for managing pulmonary disease caused by Kansasii branch mycobacteria. In cases of emergent rifamycins antimicrobial resistance (often linked to rpoB mutations[27]), substitution with alternative agents demonstrating microbiological susceptibility is recommended. Data in Shandong region reveal elevated in vitro susceptibility profiles of *M. kansasii* to rifamycins. Companion agents, including clarithromycin, amikacin, linezolid, and moxifloxacin, maintain clinically relevant susceptibility indices, positioning these agents as viable components for multidrug therapeutic protocols.

When compared to pulmonary disease caused by MAC, MABC, and *M. kansasii*, NTM-PD cases attributable to *M. fortuitum* demonstrate reduced prevalence. Antimicrobial susceptibility studies from distinct geographical regions reveal notable variations in resistance patterns. Israeli data demonstrated susceptibility profiles of ≥ 80% for amikacin, moxifloxacin, ciprofloxacin, and SXT. Conversely, clarithromycin susceptibility declined to 28%, imipenem to 56%, and tobramycin susceptibility reached a critical low of 1%[19]. Parallel analysis of British microbial isolates showed preserved efficacy (≥ 80% susceptibility) for amikacin, clarithromycin, ciprofloxacin, tigecycline, moxifloxacin, and meropenem. Notably reduced susceptibility was observed for cefotiam (49% susceptibility) and linezolid (6.9% susceptibility)[23]. This investigation identified a limited cohort of 11 *M. fortuitum-PD* cases. Comparative analysis of antimicrobial susceptibility profiles revealed variations from Israeli and British surveillance data. Linezolid and ceftriaxone demonstrated complete susceptibility (100%), followed by amikacin and moxifloxacin (90.9%), with clarithromycin exhibiting 81.8% susceptibility. Notably, doxycycline, imipenem, and tobramycin showed markedly reduced efficacy (<10% susceptibility), concurrent with elevated resistance patterns across therapeutic agents. According to the guidelines of recommendation, antimicrobial therapy for *M. fortuitum*–PD should involve a combination regimen of two to three agents. Intravenous therapeutic options include amikacin, imipenem, and cefotiam. Oral agents comprise quinolones, linezolid, according to the guidelines of recommendation, antimicrobial therapy for *M. fortuitum*–PD should involve a combination regimen of two to three agents., doxycycline, and clofazimine. Antimicrobial selection should be guided by in vitro susceptibility testing results[7]. According to the antimicrobial susceptibility profile observed in this investigation, empirical therapeutic regimens in this geographical area should exclude imipenem, doxycycline, and sulfamethoxazole due to their elevated resistance rates. Conversely, antimicrobial agents demonstrating favorable susceptibility profiles, including amikacin, clarithromycin, linezolid, moxifloxacin, and ceftriaxone, may warrant clinical consideration as alternative therapeutic options. Antimicrobial susceptibility profiling of antimicrobial combinations for *M. fortuitum* isolates was precluded by insufficient specimen quantity in this study.

In accordance with international guidelines, the diagnosis of NTM-PD relies on the identification of NTM isolates in sputum or bronchoalveolar lavage fluid. Given the prolonged culture duration and the > 1-month timeframe for AST results, guideline-recommended standardized regimens are often implemented. Under such circumstances, treatment can be administered in line with the standardized treatment protocols recommended by the guidelines. Nevertheless, given that most treatment regimens involve multi-drug combinations, adverse reactions are virtually inevitable. These may include drug allergies, cardiotoxicity, hepatic impairment, and cytopenia, among others. Such complications can potentially impede the implementation of the multi-drug combination treatment plans. A retrospective study conducted in Anhui, China, compared the standardized treatment approach advocated by the guidelines with the treatment strategy based on AST for NTM-PD. The findings indicated no significant difference in therapeutic efficacy between the two approaches[29]. Our present study offers multi-drug combination susceptibility profiles based on AST for pulmonary diseases of MAC and MABC. These profiles can serve as a valuable supplement to the standardized treatment protocols. It is important to acknowledge the limitations of the Anhui study. It was a single-center investigation with a relatively small sample size of merely 101 cases and a relatively short observation period of nine months. Expanding the sample size and extending the observation period to over one year would likely enhance the study’s clinical guidance value and provide more robust evidence to strengthen clinical applicability.

This investigation evaluated pharmacological interrelationships among congeneric therapeutic agents, including clarithromycin-azithromycin and amikacin-tobramycin combinations as well as other drug pairs in MAC and MABC. Within MAC isolates, weak correlations and poor agreement were observed for clarithromycin-azithromycin, amikacin-tobramycin, and moxifloxacin-gatifloxacin pairs. While rifampicin-rifapentine demonstrated moderate correlation, their clinical agreement remained unsatisfactory. Contrastingly, *M. abscessus* isolates exhibited significant correlations between clarithromycin-azithromycin and minocycline-doxycycline combinations, whereas amikacin-tobramycin showed no detectable association. Three potential explanations emerge: 1. Drug-class-specific resistance mechanisms may extend beyond currently characterized pathways, suggesting individualized resistance patterns. 2. Current susceptibility breakpoint determinations appear compromised by insufficient high-quality clinical trial data and retrospective analyses, introducing potential systematic bias. The evidence gap in pharmacological validation underscores the imperative for controlled clinical studies to establish reliable susceptibility criteria.

Nontuberculous mycobacteria are classified as rapid- or slow-growing mycobacteria based on their growth rates. The rapid-growing group comprises *M. abscessus*, *M. fortuitum*, *Mycobacterium smegmatis*, and *M. chelonae*, among others. Slowly growing species include MAC, *M. xenopi*, *M. kansasii*, *Mycobacterium simiae*, *Mycobacterium scrofulaceum*, and *Mycobacterium szulgai*, among others. This study revealed distinct antimicrobial susceptibility patterns between slow - growing mycobacteria (predominantly MAC and *M. kansasii*) and rapid growing mycobacteria (primarily M. abscessus, *M. fortuitum and M. chelonae*). Both MAC and *M. kansasii* demonstrated comparable susceptibility profiles for the most effective agents, including clarithromycin, amikacin, rifamycins, and moxifloxacin. However, ethambutol and gatifloxacin exhibited notably low susceptibility rates with elevated resistance in MAC isolates, whereas these agents maintained higher efficacy against *M. kansasii*. Some studies have demonstrated that isoniazid is effective for the treatment of *M. kansasii*, yet exhibits lower activity in the treatment of MAC. Among rapid-growing mycobacteria, susceptibility rates remained consistently elevated for clarithromycin, moxifloxacin, cefoxitin, amikacin, and linezolid. Due to the intrinsic resistance of rapid-growing mycobacteria to rifamycins, susceptibility testing for this drug class was omitted. Significant variability emerged in susceptibility profiles for other antimicrobials, particularly azithromycin and ethambutol, which demonstrated inconsistent activity patterns across tested isolates. This investigation conducted a systematic analysis of antimicrobial susceptibility profiles across differential growth rates of NTM isolates, revealing marked variations between distinct mycobacterial subgroups. The antimicrobial resistance patterns in NTM demonstrate significant correlations with both growth kinetics and phylogenetic divergence, thereby underlying the divergent therapeutic approaches required for pulmonary infections caused by rapidly growing versus slow-growing NTM species.

While macrolides, amikacin, and rifampin exhibit in vitro susceptibility profiles that correlate with therapeutic outcomes in specific nontuberculous mycobacterial pulmonary diseases — including MAC-PD, *M. kansasii*-PD, and MABC-PD, the clinical significance of susceptibility testing results for other antimicrobial agents remains unsubstantiated. This uncertainty persists due to insufficient validation through large-scale clinical trials establishing laboratory-to-clinic efficacy correlations. Current evidence demonstrates significant geographic heterogeneity in antimicrobial susceptibility patterns among NTM species. When developing treatment regimens based on authoritative guidelines, it is important to note that, except for *M. kansasii*-PD, the treatment failure rates for MAC-PD and MABC-PD are relatively high. Therefore, When formulating therapeutic regimens, clinical decision-making must be guided not only by consultation of evidence-based clinical guidelines but also by incorporation of antimicrobial susceptibility testing data. Current research demonstrates a limited correlation between in vitro antimicrobial susceptibility profiles and clinical effectiveness; nevertheless, pharmaceutical agents demonstrating phenotypic resistance through susceptibility testing methodologies frequently exhibit suboptimal therapeutic performance in clinical settings. Such agents should be excluded from empirical treatment protocols unless rigorous pharmacological investigations establish verifiable synergistic interactions between therapeutic compounds. The ATS guidelines endorse amikacin combination therapy with intravenous agents for rapid-growth mycobacterial pulmonary infections, specifying imipenem as the preferred agent. However, our regional surveillance data from Shandong Province demonstrate markedly elevated resistance rates to imipenem among rapid-growth mycobacterial isolates, rendering this agent unsuitable for empirical therapeutic regimens in this geographical context. For MAC pulmonary disease management, susceptibility patterns in Shandong contrast with guideline recommendations. While azithromycin is endorsed in the ATS guidelines based on in vitro susceptibility data, regional evidence demonstrates clarithromycin achieves superior susceptibility rates compared to azithromycin. This warrants prioritization of clarithromycin as the core therapeutic agent for MAC disease in local empirical regimens. Although ethambutol demonstrates relatively low sensitivity rates in our region (clarithromycin 98.75% versus azithromycin 30.25%; P < 0.01), its inclusion in treatment protocols remains justified by evidence showing reduced macrolide resistance emergence[20]. Current clinical practice should retain ethambutol as an adjunctive component until updated resistance surveillance data become available. Certain antimicrobial agents exhibit in vitro activity in drug susceptibility assays, yet demonstrate elevated minimum inhibitory concentration (MIC) values. Achieving these MIC thresholds in vitro necessitates the administration of excessively high dosages, which correlate with an elevated incidence and severity of adverse effects. For RGM, CLSI guideline M62 ED1 establishes linezolid susceptibility breakpoints at 8-32 µg/ml. While a strain demonstrating a linezolid MIC of 8 µg/ml meets susceptibility criteria, pharmacokinetic analysis reveals that attaining therapeutic concentrations would necessitate supraphysiological dosing in clinical practice, potentially inducing severe adverse effects. This underscores the necessity for comprehensive therapeutic evaluation integrating both antimicrobial susceptibility data and quantitative MIC values. Clinical decision-making should incorporate MIC-guided dose optimization to balance microbiological efficacy with pharmacological safety parameters.

## Conclusion

Given the distinct drug resistance profile of nontuberculous mycobacteria (NTM) in Shandong Province, establishing a region-specific empirical treatment protocol based on guideline-recommended drug combinations has become imperative. Concurrently, prospective cohort studies should be conducted to investigate the correlation between antimicrobial susceptibility testing (AST) results and clinical outcomes, thereby enhancing the clinical utility of AST in therapeutic decision-making. With advancements in technology and improvements in diagnostic capabilities among healthcare professionals, the isolation rate of non-tuberculous mycobacteria (NTM) and the diagnostic rate of NTM-related pulmonary disease (NTM-PD) have been steadily increasing. Global management of NTM-PD continues to encounter substantial obstacles, including diagnostic confusion with tuberculosis, inappropriate therapeutic interventions, and elevated rates of unsuccessful treatments. Significant variations in pathogenic species distribution and antimicrobial susceptibility profiles across geographical regions further complicate clinical decision-making. Current therapeutic strategies face dual limitations: rigid adherence to clinical guidelines may prove ineffective given microbial diversity, while antimicrobial susceptibility testing (AST)-guided drug selection encounters practical implementation barriers. The absence of multinational clinical trials and slow-paced antimicrobial development have prevented establishment of standardized treatment protocols comparable to tuberculosis management paradigms. While there is insufficient evidence to establish a correlation between the results of NTM-PD susceptibility testing for certain drugs and their clinical efficacy, AST continues to play a pivotal role in guiding therapeutic regimen development. This methodology will further provide essential reference parameters for forthcoming clinical investigations. Prioritization of standardized AST implementation is therefore imperative, requiring evidence-based selection of antimicrobial agents for testing panels and establishment of clinically relevant breakpoints through rigorous clinical validation studies.

## Data Availability

All relevant data are within the manuscript and its Supporting Information files.

## Acknowledgements

We sincerely acknowledge the colleagues from our department and the Laboratory Medicine Division of Shandong Public Health Clinical Center for their professional and technical support throughout this study. Their expertise and dedication were invaluable to the completion of this research.

